# Time to SARS-CoV-2 Clearance Among Patients with Cancer and COVID-19

**DOI:** 10.1101/2020.07.23.20161000

**Authors:** Wenxin Xu, Andrew J. Piper-Vallillo, Poorva Bindal, Jonathan Wischhusen, Jaymin M. Patel, Daniel B Costa, Mary Linton B. Peters

**Author notes:** **Address correspondence to:** Mary Linton B Peters, MD, Beth Israel Deaconess Medical Center, 330 Brookline Ave, Shapiro 913, Boston, MA 02215-5400. contributed equally to this work (Wenxin Xu, Andrew J. Piper-Vallillo, Poorva Bindal, Jonathan Wischhusen).

## Abstract

**Key Points:** *Question:* What is the median time to clearance of SARS-CoV-2 among cancer patients according to currently used criteria?

*Findings:* In this single-institution retrospective cohort study, the median time to SARS-CoV-2 clearance was 50 days using the ASCO/CDC criteria of 2 negative RT-PCR assays >24 hours apart. Using alternative criteria of 1 negative RT-PCR assay (UK-NICE) or CDC clinical criteria (10 days after first positive RT-PCR and 3 days after last symptoms), median clearance times were 31 days and 13 days, respectively.

*Meaning:* SARS-CoV-2 clearance times differ substantially depending on criteria used and may be prolonged in cancer patients.

## Background

COVID-19 in oncologic patients presents a clinical dilemma given the competing priorities of providing timely anti-cancer therapy, avoiding immunosuppression during COVID-19 infection, and minimizing exposure risk to others. Several different criteria for COVID-19 clearance have been proposed.^1–3^ However, the time to clearance for cancer patients under each of these criteria is not established.

## Methods

We identified all patients at a tertiary care hospital between March 25, 2020 and June 6, 2020 with a positive nasopharyngeal SARS-CoV-2 RT-PCR, a cancer-related clinical visit within 3 years, and at least one follow-up SARS-CoV-2 assay. Data collection was performed as part of an institutional COVID-19 registry project and approved by the institutional review board.

SARS-CoV-2 RT-PCR testing was performed using the Abbott Laboratories m2000 platform in conjunction with either the Aldatu Biosciences PANDAA qDxTM SARS-CoV-2 or Abbott RealTime SARS-CoV-2 assays. At our institution, and per current ASCO guidelines, discontinuation of COVID-19 precautions requires two consecutive negative PCR tests >24 hours apart. Time to COVID-19 clearance was analyzed using the Kaplan-Meier probability estimator, for which data was censored at the time of the last known COVID-19 RT-PCR assay.

## Results

We identified 32 patients who met inclusion criteria (**Supplementary Figure S1**). Patient and tumor characteristics are described in **Table 1**. 16 patients had metastatic disease at the time of COVID-19 diagnosis. 17 patients were on active treatment at the time of COVID-19 diagnosis, 8 receiving cytotoxic chemotherapy.

**Table 1:** Baseline patient characteristics Median Age

|  |  |
| --- | --- |
| **Median Age** | 65 |
| **Sex** |  |
| Male | 14 |
| Female | 18 |
| **Race** |  |
| White | 15 |
| Black | 7 |
| Asian | 2 |
| Other/Hispanic | 4 |
| Unknown | 4 |
| **Alcohol Use** |  |
| Yes - Current | 8 |
| Yes - Past | 3 |
| No/Unknown | 21 |
| **Tobacco Use** |  |
| Yes - Current | 2 |
| Yes - Past | 11 |
| No/Unknown | 19 |
| **Cardiac Disease (CAD, MI, CHF, or Arrhythmia)** |  |
| Yes | 11 |
| **Hypertension** |  |
| Yes | 16 |
| **Diabetes Mellitus, Type 2** |  |
| Yes | 7 |
| **Presenting Symptoms** |  |
| Symptomatic | 27 |
| Asymptomatic | 5 |
| **ECOG PS** |  |
| 0-I | 24 |
| II | 4 |
| III | 1 |
| IV | 1 |
| Unknown | 2 |
| **Pre-COVID Treatment Characteristics** |  |
| Received Treatment within 6 months of diagnosis | 17 |
| Received Chemotherapy | 7 |
| Received Immunotherapy | 2 |
| Received Targeted therapy | 6 |
| Received Hormonal therapy | 3 |
| **Tumor Type** |  |
| Gastrointestinal tumors | 9 |
| Hematologic malignancies | 7 |
| Breast and gynecologic tumors | 6 |
| Genitourinary tumors | 5 |
| Thoracic tumors | 3 |
| Other* | 2 |

\* Includes one patient with melanoma and one patient with osteosarcoma

Using our institutional criteria, 13 patients were cleared of COVID-19 precautions, at a median time of 33 days (range 10-58 days) (**Figure 1**). Across the full cohort, median time to clearance was estimated at 50 days (95% CI, 33-58 days) (**Figure 2**). 14 patients resumed anti-cancer treatment prior to clearance, requiring substantial allocation of resources for isolation during treatment. All patients were asymptomatic from COVID-19 at the time of resuming anti-cancer treatment, and experienced no subsequent complications attributable to COVID-19. Among the 13 patients who met clearance criteria, 2 subsequently had a positive RT-PCR test.

**Figure 1:**
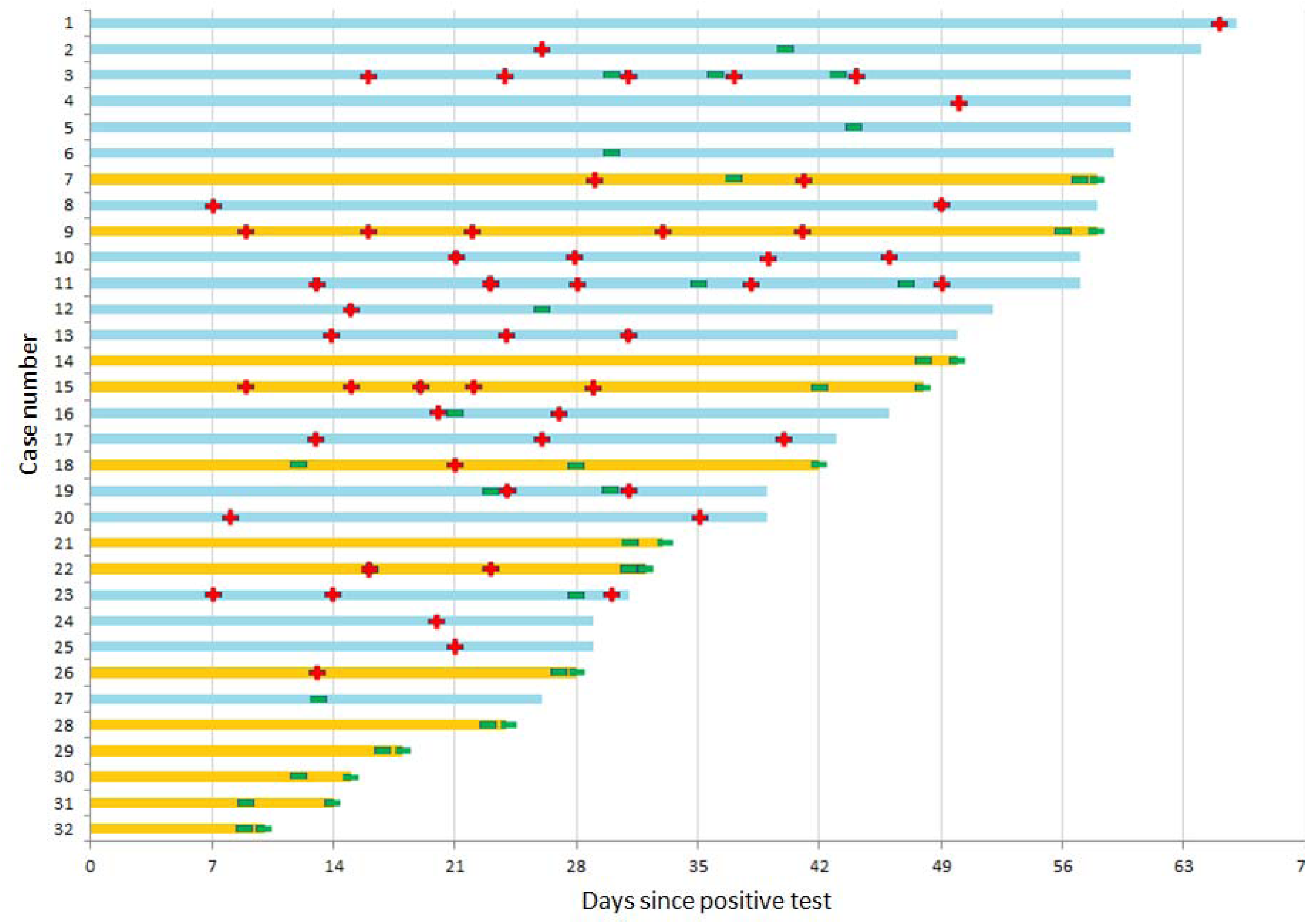
Swimmer’s plot of COVID-19 testing results. (+) indicates SARS-CoV-2 positive RT-PCR from a nasopharyngeal specimen. (-) indicates SARS-CoV-2 negative RT-PCR. Orange bars indicate patients who were cleared during the follow-up period based on 2 negative RT-PCR results >24 hours apart. Patients #21 and #26 later had a subsequent positive SARS-CoV-2 PCR after meeting clearance criteria.

**Figure 2:**
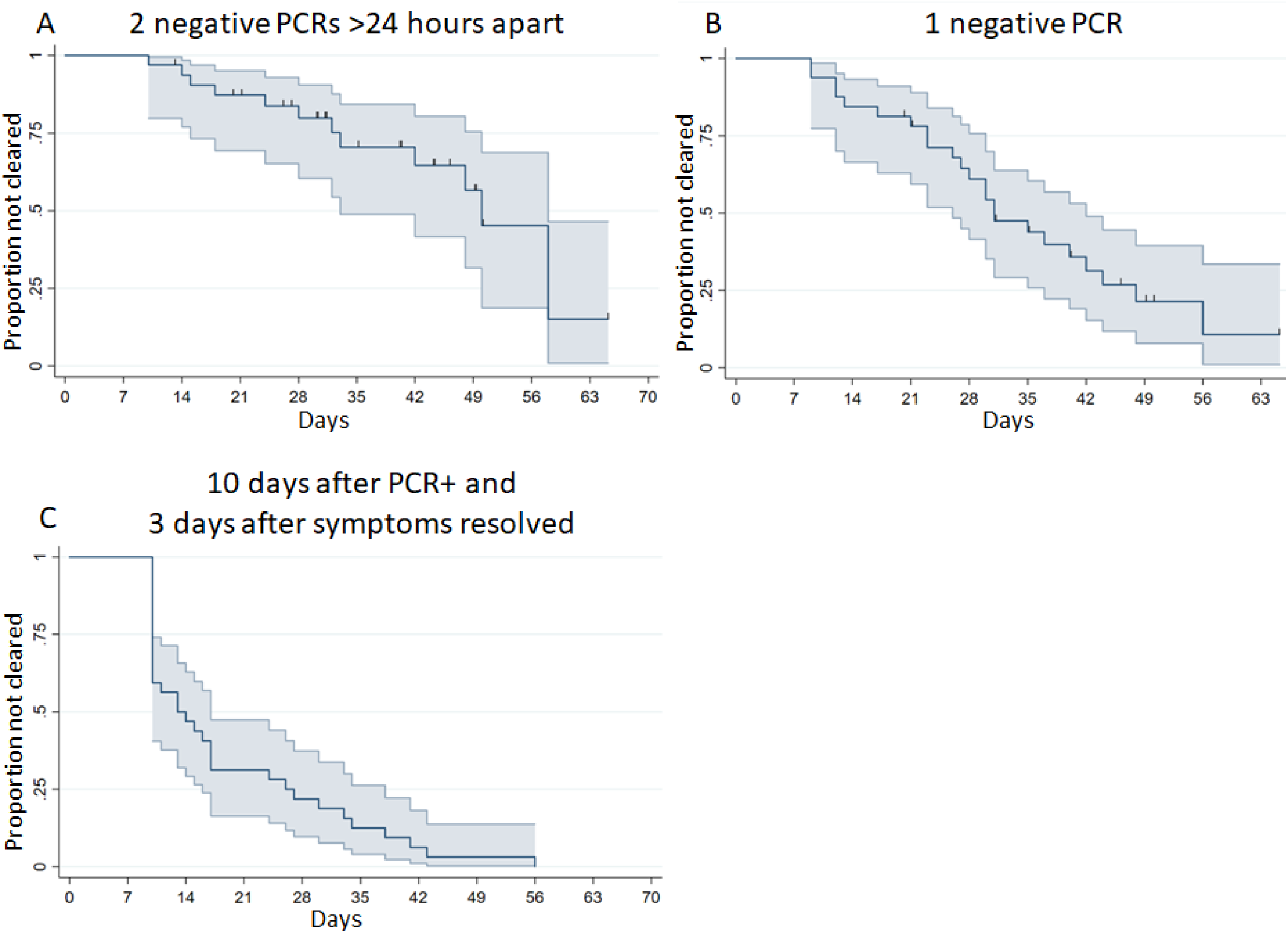
Time to COVID-19 clearance under different criteria. Kaplan-Meier probability estimate functions are shown for three alternative criteria for defining COVID-19 precaution clearance: A) ASCO/CDC test-based guideline of 2 negative RT-PCR results > 24 hours apart, B) UK-NICE guideline of 1 negative RT-PCR after the initial COVID-19 diagnosis, and C) revised CDC symptom-based recommendations for clearance. In panels A and B, patients are censored at time of last SARS-CoV-2 PCR assay. In panel C but no censoring was needed as all patients achieved clearance at last clinical follow-up.

We calculated COVID-19 clearance times under alternative criteria. Using the UK-NICE guidelines (one negative RT-PCR test), median time to clearance would have been 31 days (95% CI, 26-42 days). Using a symptom/time-based strategy per CDC criteria (10 days after first positive RT-PCR and 3 days after last symptoms), median time to clearance would have been 13 days (95% CI, 10-17 days) (**Figure 2**). Cox proportional hazards models showed that patients with symptoms from COVID-19 versus those asymptomatically detected had longer time to first negative RT-PCR (HR 0.25 for negative PCR, 95% CI 0.08-0.76, p=0.01). No significant associations were seen between anti-cancer treatment before or during COVID-19 positivity and clearance time (data not shown).

## Discussion

In this study, we observe that time to nasopharyngeal SARS-CoV-2 RNA clearance in our oncology patients is substantially longer than the approximately 17-20 days previously reported in the general population.^4–6^ The calculated time to COVID-19 clearance varies widely under the three clinical criteria examined. Choosing criteria for COVID-19 clearance in cancer patients must take into account the tradeoffs between the increased risk of poor clinical outcomes from COVID-19, the clinical risk from delays in anti-cancer therapy, and the risk of exposing others.^7,8^ In addition, further elucidation of the relationship between viral shedding and infectivity is needed to guide the development of future guidelines.

## Conclusions

In patients with cancer who develop COVID-19 infection, viral shedding can persist for many weeks after disease onset. Symptom/time-based strategies result in shorter times to discontinuation of precautions and resumption of treatment compared to test-based strategies, though the risks of earlier return to routine cancer care remain unclear.

## Data Availability

Blinded data available for review on request

## Acknowledgments

We would like to acknowledge the clinical staff, nurses, fellows and faculty assigned to our COVID-19 outpatient/inpatient unit (HO-REEES), and our patients with a diagnosis of cancer that were afflicted during the COVID-19 pandemic.

## Disclosure of Potential Conflicts of Interest

### JMP

Received honoraria from Radius, research grant support from Breast Cancer Research Foundation, institutional research support from Genentech, Sanofi, Odonate Therapeutics, all outside submitted work.

### DBC

Reports personal fees (consulting fees and honoraria) and nonfinancial support (institutional research support) from Takeda/Millennium Pharmaceuticals, and AstraZeneca, and Pfizer, as well as nonfinancial support (institutional research support) from Merck Sharp and Dohme Corporation, Merrimack Pharmaceuticals, Bristol-Myers Squibb, Clovis Oncology, Spectrum Pharmaceuticals and Tesaro, all outside the submitted work.

### MLBP

Received honoraria from Bayer, Exelixis and Agios, travel support from Halozyme, AstraZeneca and Exelixis, research support from Bayer, institutional research support from Taiho, AstraZeneca, BeiGene, Berg, Merck, all outside the submitted work.

---

**Supplemental Figure S2:**
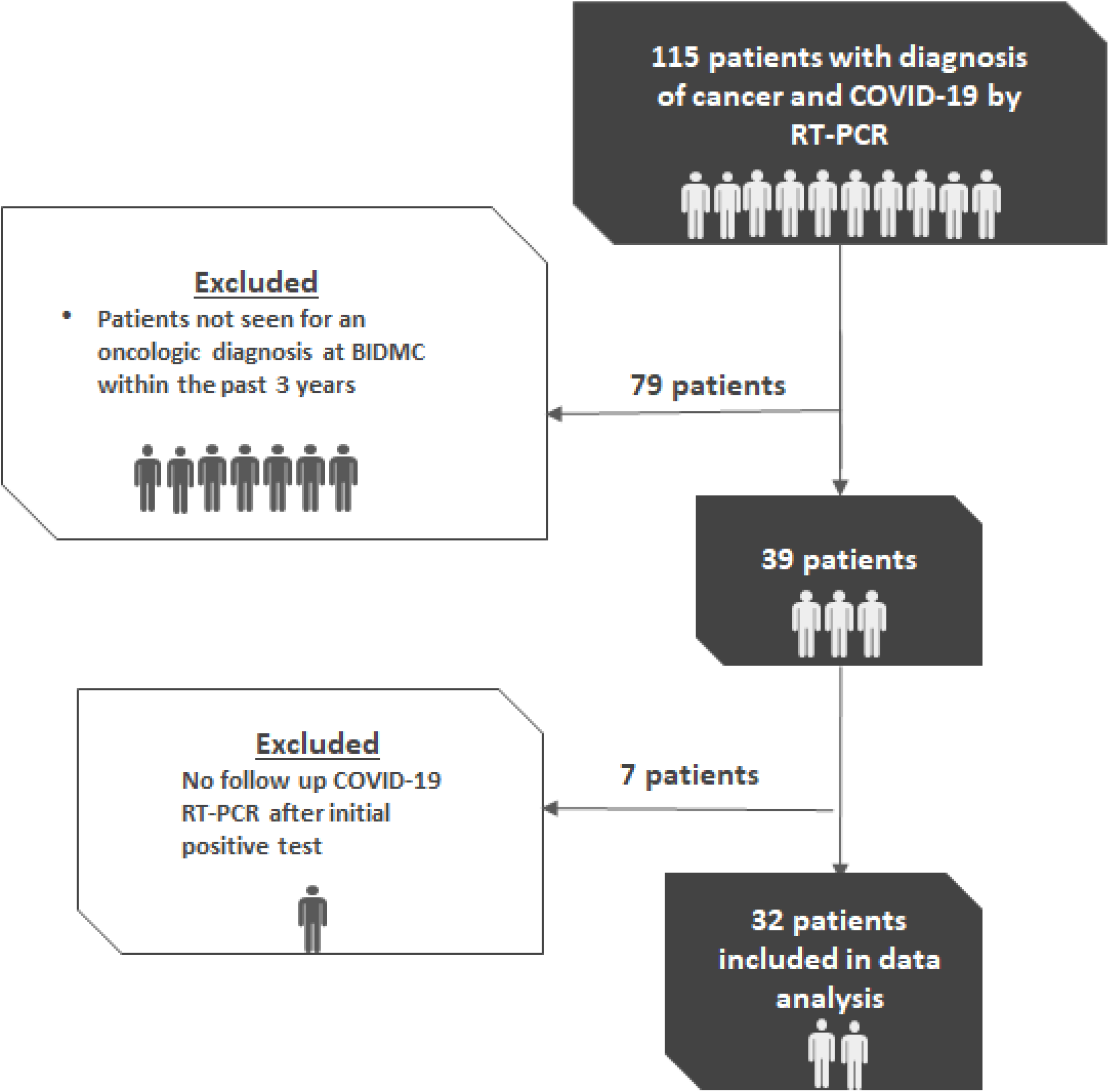
CONSORT diagram showing cohort selection.

## REFERENCES

1. Centers for Disease Control and Prevention. Coronavirus Disease 2019 (COVID-19). Centers for Disease Control and Prevention. Published February 11, 2020. Accessed June 18, 2020. https://www.cdc.gov/coronavirus/2019-ncov/hcp/disposition-hospitalized-patients.html

2. American Society of Clinical Oncology COVID-19 Advisory Group. COVID-19 Patient Care Information. Accessed June 18, 2020. https://www.asco.org/asco-coronavirus-information/care-individuals-cancer-during-covid-19

3. The National Institute for Health and Care Excellence. COVID-19 rapid guideline: delivery of systemic anticancer treatments. Accessed June 18, 2020. https://www.nice.org.uk/guidance/ng161

4. Xu K, Chen Y, Yuan J, et al. Factors associated with prolonged viral RNA shedding in patients with COVID-19. Clin Infect Dis. Published online April 9, 2020. doi:10.1093/cid/ciaa351

5. Zhou F, Yu T, Du R, et al. Clinical course and risk factors for mortality of adult inpatients with COVID-19 in Wuhan, China: a retrospective cohort study. The Lancet. Published online March 2020:S0140673620305663. doi:10.1016/S0140-6736(20)30566-3

6. Zheng S, Fan J, Yu F, et al. Viral load dynamics and disease severity in patients infected with SARS-CoV-2 in Zhejiang province, China, January-March 2020: retrospective cohort study. BMJ. 2020;369:m1443. doi:10.1136/bmj.m1443

7. Liang W, Guan W, Chen R, et al. Cancer patients in SARS-CoV-2 infection: a nationwide analysis in China. The Lancet Oncology. 2020;21(3):335–337. doi:10.1016/S1470-2045(20)30096-6

8. Kuderer NM, Choueiri TK, Shah DP, et al. Clinical impact of COVID-19 on patients with cancer (CCC19): a cohort study. The Lancet. Published online May 2020:S0140673620311879. doi:10.1016/S0140-6736(20)31187-9

